# Resting-state fMRI Connectivity between Semantic and Phonologic Regions of Interest May Inform Language Targets in Aphasia

**DOI:** 10.1101/2020.06.29.20142786

**Authors:** Amy E. Ramage, Semra Aytur, Kirrie J. Ballard

## Abstract

**Purpose:** Brain imaging has provided puzzle pieces in the understanding of language. In neurologically healthy populations, structure of certain brain regions is associated with particular language functions (e.g., semantics, phonology). In studies on focal brain damage, certain brain regions or connections are considered sufficient or necessary for a given language function. However, few of these account for the effects of lesioned tissue on the *functional* dynamics of the brain for language processing. Here, functional connectivity amongst semantic-phonologic regions of interest (ROIs) is assessed to fill a gap in our understanding about the neural substrates of impaired language and whether connectivity strength can predict language performance on a clinical tool in individuals with aphasia.

**Method:** Clinical assessment of language, using the *Western Aphasia Battery-Revised* (WAB**-**R), and resting-state fMRI data were obtained for 30 individuals with chronic aphasia secondary to left hemisphere stroke and 18 age-matched healthy controls. Functional connectivity (FC) between bilateral ROIs was contrasted by group and used to predict WAB-R scores.

**Results:** Network coherence was observed in healthy controls and participants with stroke. The left-right premotor cortex connection was stronger in healthy controls, as reported by New et al. (2015) in the same data set. FC of (1) bilateral connections between temporal regions, in the left hemisphere and bilaterally, predicted lexical semantic processing for Auditory Comprehension and (2) ipsilateral connections between temporal and frontal regions in both hemispheres predicted access to semantic-phonologic representations and processing for verbal production.

**Conclusions:** Network connectivity of brain regions associated with semantic-phonologic processing is predictive of language performance in post-stroke aphasia. The most predictive connections involved right hemisphere ROIs – particularly those for which structural adaptions are known to associate with recovered word retrieval performance. Predictions may be made, based on these findings, about which connections have potential as targets for neuroplastic functional changes with intervention in aphasia.

---

Several modern brain imaging methodologies have been used to investigate aphasia, each providing its piece in understanding language processing. The tradition of attaching cognitive function to particular brain regions began long before imaging was available as a tool, when Paul Broca and Carl Wernicke made their postmortem discoveries. Broca provided the first post-mortem evidence of language function localization, i.e., that the ability to articulate language is in the posterior 2/3 of the inferior frontal gyrus (Grodzinsky & Amunts, 2006). About 10 years later, Wernicke found that damage to the superior temporal gyrus is associated with language comprehension. This established brain-behavior relationships, or functional specialization of brain regions (Geschwind, 1970). With the advent of neuroimaging, the mapping of language functions to brain regions has become more complex (Holland & Ramage, 2000), facilitating a narrowing of the focus to specific language functions.

Current neuroimaging data hinders translation of research to practice given the multitude of approaches taken to identify the brain regions that are necessary or sufficient for language processing (Poldrack, 2000; Price et al., 1999; Price & Mummery, 1999). These methods often yield different results regarding which regions and functions are most important for studies of language or aphasia, largely because the methodology or population of study differs across studies. For example, some studies identify regions that are necessary for processing of word meanings, or semantics (e.g., Halai et al., 2017; Jackson et al., 2016), or for phonology (e.g., Okada, Matchin, & Hickok, 2018) in neurologically healthy populations using task-based functional MRI (fMRI). Others study individuals with focal brain damage to indicate which brain *regions* (Price, 2012) or structural *connections* (Fridriksson et al., 2018; Ivanova et al., 2016) are necessary or sufficient for a given language function. However, few investigate the causal relationship between a brain lesion to an impaired function to indicate the *necessity* of a region for a specific function (Poldrack, 2000).

## Lesion Studies

While isolating language processes for study may not be representative of language as the dynamic process that it is, this gold standard approach has yielded immense understanding about language and the brain. This approach is exemplified by Price and colleagues, who have developed a cumulative model of language processes and their associated brain regions (Price, 2012). They have identified lesions that are necessary to disrupt specific language functions in persons with aphasia secondary to left hemisphere stroke (PWAs) (Hope et al., 2017; Price, 2012; Seghier et al., 2010) establishing specific language process-to-brain lesion correspondence (Seghier et al., 2015) and improving understanding of recovery from aphasia. However, most of the predictions have been tested as they relate to language performance during task-based fMRI in healthy, neurologically normal adults (e.g., Hope et al., 2014; Ludersdorfer et al., 2019). Only a few have been tested using fMRI in individuals with aphasia, as described below.

Similarly, the work of Schwartz and colleagues has focused on isolating components of language processing to associate them with lesion characteristics. This work has also carefully characterized language using single word processing assessments to specify semantics (Schnur et al., 2009; Schwartz et al., 2009), phonology (Schwartz et al., 2012; Thompson-Schill et al., 2010), and sentence comprehension and production (Thothathiri et al., 2010, 2015; Thothathiri, Gagliardi, et al., 2012; Thothathiri, Kimberg, et al., 2012). These studies have utilized voxel-based lesion symptom mapping (VLSM) which associates lesions in particular brain areas (defined by voxels) with language symptoms and therefore the results are relevant to lesion locations present in the participant sample and do not assess the dynamic functional brain activity.

## Structural Connectivity Studies

As such, there remains a gap in knowledge about whether or how well previous findings based on structural, anatomical or lesion-based studies are suited for selecting brain regions that may be informative in *functional* imaging studies to understand language breakdown in individuals with aphasia. That is, lesion studies indicate the areas of the brain that are *necessary* to perform a language process, and activation studies in neurotypical individuals indicate brain regions that are *sufficient* to perform a language process, but neither indicates how those regions interact to sufficiently approximate language in aphasia. It is likely that, rather than only investigating structural characteristics of lesions within certain brain regions, investigations of structural or functional connections amongst regions (i.e., connectivity) may facilitate prediction of aspects of language performance in individuals with aphasia (Pustina et al., 2017).

Voxel-based and connectivity-based lesion-symptom mapping (e.g., den Ouden et al., 2019; Fridriksson et al., 2018) and metrics of white matter integrity (e.g., Ivanova et al., 2016) have been used to correlate or predict language performance in individuals with aphasia. Language performance in these studies has been characterized using aphasia batteries (e.g., *Western Aphasia Battery-Revised*, WAB-R, (Kertesz, 2007)) as well as with assessments of semantic associations (e.g., with the Pyramids and Palm Trees test, Howard & Patterson, 1992) and other word retrieval measures (e.g., the Philadelphia Naming Test (Roache et al., 1996) that includes items that differ by word class) that are used in clinical practice. These studies have improved understanding of the structural connections most likely involved in language processing, and have even documented changes in white matter integrity as a function of treatment (McKinnon et al., 2017). They have also informed the contribution of the right hemisphere in recovery in aphasia, with structural adaptation that is uniquely associated with recovery of specific language performance (Hope et al., 2017). However, a gap remains in understanding how lesions affecting white matter integrity or connectivity change the functional dynamics of the brain for language performance in aphasia. In fact, recent predictive models of language performance indicate that structural connectivity metrics are highly correlated with lesion volume and thus do not add much to the functional predictions about language performance (Hope et al., 2017).

## Functional Connectivity Studies

There have been several informative studies of the functional dynamics of brain function in aphasia with fMRI using language tasks (e.g., Kiran & Thompson, 2019) or by modeling the causal dynamics of task-based fMRI (e.g., Meier, Kapse, & Kiran, 2016). However, as noted by Klingbeil and colleagues (Klingbeil et al., 2017), performing language tasks in an MRI scanner is awkward at best, and performance under such conditions is dubious given the known intra- and inter-individual variability that is characteristic of individuals with aphasia. There continues to be a need to identify relatively coherent brain states, that are impervious to differing task demands, and that may be investigated (1) in recovery from aphasia and (2) as targets to assess neuroplastic change associated with a treatment.

Resting-state fMRI (rsfMRI) provides information about brain networks that demonstrate intrinsic coherent activity. Networks demonstrating intrinsic coherent activity at rest are impervious to task-related fluctuations and are considered to be more permanent, trait-like functional signatures (Hjelmervik et al., 2014). In fact, resting-state intrinsic networks are thought to provide the scaffolding for activity engaged during task performance (Chai et al., 2016; Smith et al., 2009). For example, Jackson and colleagues (Jackson et al., 2016) investigated a network of regions functionally connected to the ventral anterior temporal gyrus in rsfMRI to establish a ‘semantic network’, based on the premise that this brain region is a critical hub of multimodal semantic conceptual representations. They then compared that network at rest with patterns of activation during a triad semantic judgement task (e.g., matching a target word – ‘hen’ – to either a strongly associated word – ‘cage’ – or conceptually similar word – ‘robin’ – relative to unrelated foils), further specifying the components of the network to semantic processing. Thus, resting-state fMRI can index intrinsic connectivity networks that may or may not be anatomically connected, but that have relevance to the networks that are engaged during language performance. In concert with the exquisite detail provided by studies of post-stroke lesions and structural connectivity (Fridriksson et al., 2018; Halai et al., 2018; Hope et al., 2017; Price, 2012; Seghier et al., 2015), and the process-based focus of task-based fMRI studies (Hope et al., 2014), resting-state fMRI is a complementary tool providing a simple yet robust indicator of network structure that may be useful in studying the relationships of these networks to performance.

Thus, the present study utilizes rsfMRI to determine whether a set of regions found previously to be *necessary* for semantic or phonologic processing (Halai, Woollams, & Lambon Ralph, 2017; Seghier et al., 2010) or for speech motor planning (New et al., 2015; Price, 2012) demonstrate coherent activity (a network) at rest in healthy controls and PWAs. The *a priori* hypothesis underlying the study is that coherent activity between regions of interest present in the Control group represents a resting-state intrinsic network that serves as the scaffolding upon which language performance depends (Fox & Raichle, 2007; Smith et al., 2009). *Group differences* in functional connectivity strength (FC) amongst this network between PWAs and control participants may have relevance for language performance; therefore, the present study assesses the *predictive value* of FC amongst these regions for language performance in aphasia using a commonly-used clinical tool – the WAB-R. While this aphasia battery does not isolate language processes specifically, it provides a unique opportunity to link regions-of-interest (ROIs) to semantic and phonologic processing data typically collected in clinical settings. Thus, we determine how well FC amongst these ROIs can predict overall aphasia severity (the WAB-R Aphasia Quotient), as well as subtest scores for Spontaneous Speech (with subcomponents Content and Fluency), Auditory Comprehension, Repetition, and Naming/Word Finding.

## Method

### Participants

An existing data set was used for the study which included thirty-one, right-handed, chronic left hemisphere stroke participants with aphasia (New et al., 2015). Speech-language assessments and rsfMRI connectivity analyses were conducted. Fifteen of the participants had comorbid apraxia of speech (see New et al., 2015, for more details). Eighteen healthy volunteers (HC) without neurological or psychiatric disorders also participated. All participants were: 18-75 years of age, right-handed, self-reported (by PWA or caregiver) proficient English speakers prior to the stroke and had no contraindications for undergoing an MRI. Additionally, eligible participants had no history of uncorrected hearing, vision, or other sensory impairments; cognitive impairments; premorbid speech, language, or reading impairments; or substance abuse. All control participants scored within normal limits (> 28) on the Mini-Mental Status Examination (MMSE; Folstein et al., 1975). All procedures conformed to the Declaration of Helsinki (BMJ 1991; 302: 1194) and were approved by the Sydney Local Health District Human Research Ethics Committee, Concord Repatriation General Hospital in Sydney and the University of Sydney, Australia. The examiner, a qualified speech-language pathologist, walked through the participant information statement and consent form with the participant and caregiver, and asked comprehension questions to ensure understanding of study details prior to obtaining written consent. All participants provided written informed consent. In addition, because the assessments included verbal expression, participants were excluded if they had apraxia of speech judged as severe, as described below.

#### Speech and Language Evaluation

Because the data used here are from an existing data set, we did not have the ability to select our own test batteries. The existing data set included stroke participants’ performance on a battery of assessments used to diagnose and determine presence and severity of dysarthria, apraxia, as well as severity and type of aphasia. The WAB-R was administered as a measure of aphasia severity and served as the primary measure of language in the study. While the WAB-R does not isolate semantics or phonology, access to both is required to a varying extent for all subtests.

Additional measures were administered to rule out auditory discrimination deficits (Psycholinguistic Assessments of Language Processing in Aphasia – Same-Different Discrimination Using Word Minimal Pairs subtest (Kay et al., 1992)) and nonverbal cognitive deficit (Raven’s Progressive Colored Matrices (Raven, 1978)). The Motor Speech Examination (Duffy, 2013) and Apraxia Battery for Adults – 2 (Dabul, 2000) – increasing word length subtest, characterized motor speech. For published tests that involve perceptual judgment and scoring (e.g., WAB-R Fluency subtest), research assistants were trained by author KJB and discrepancies in scoring were discussed and determined by consensus. Reliability for the ABA-2 Increasing Word Length, inter-rater reliability was 98%. Additionally, connected speech samples were elicited either through the Story Retell Procedure (McNeil et al., 1997) or unconstrained conversation with the examiner (Ballard et al., 2016). All testing was completed by research assistants or author KJB. A research assistant edited (to remove silences and off-task behaviors) and compiled the recorded tasks into a single ∼20 minute video, with the responses to each task labelled in sequence. These videos were uploaded to a secure server that was accessed by the three expert judges (Ballard et al., 2016). The videos contained no identifying information, other than the audiovisual image. These judges were blinded to any other demographic information, did not administer any of the testing, and were blinded to scores of any formal tests (e.g. WAB-R, ABA-2 subtests, PALPA subtest) or neuroimaging findings at the time of making their ratings. They judged participant speech samples for presence/severity of apraxia, dysarthria, nonverbal oral apraxia and phonologically-based sound errors on a 7 point Likert scale (1 = normal, 7 = severe; see (Ballard et al., 2016; New et al., 2015) for more details).

Participants’ demographic variables and individual testing performance are reported in Table 1. All participants spoke English proficiently, though there was one participant whose first language was Hindi (DIS048) and one whose first language was Cantonese (DIS006). Neither were outliers for any of the language or neuroimaging measures, and therefore were included in the study. One PWA (DIS002) was removed as he had extreme outlier values for both WAB-R AQ and AC. Figure 1 demonstrates the extent and overlap of lesions in the sample. Language performance on the WAB-R for each participant is presented in Table 2. The PWA and HC groups did not differ for age or education level, but there were more females in the HC than PWA group (Table 2).

**Table 1.**
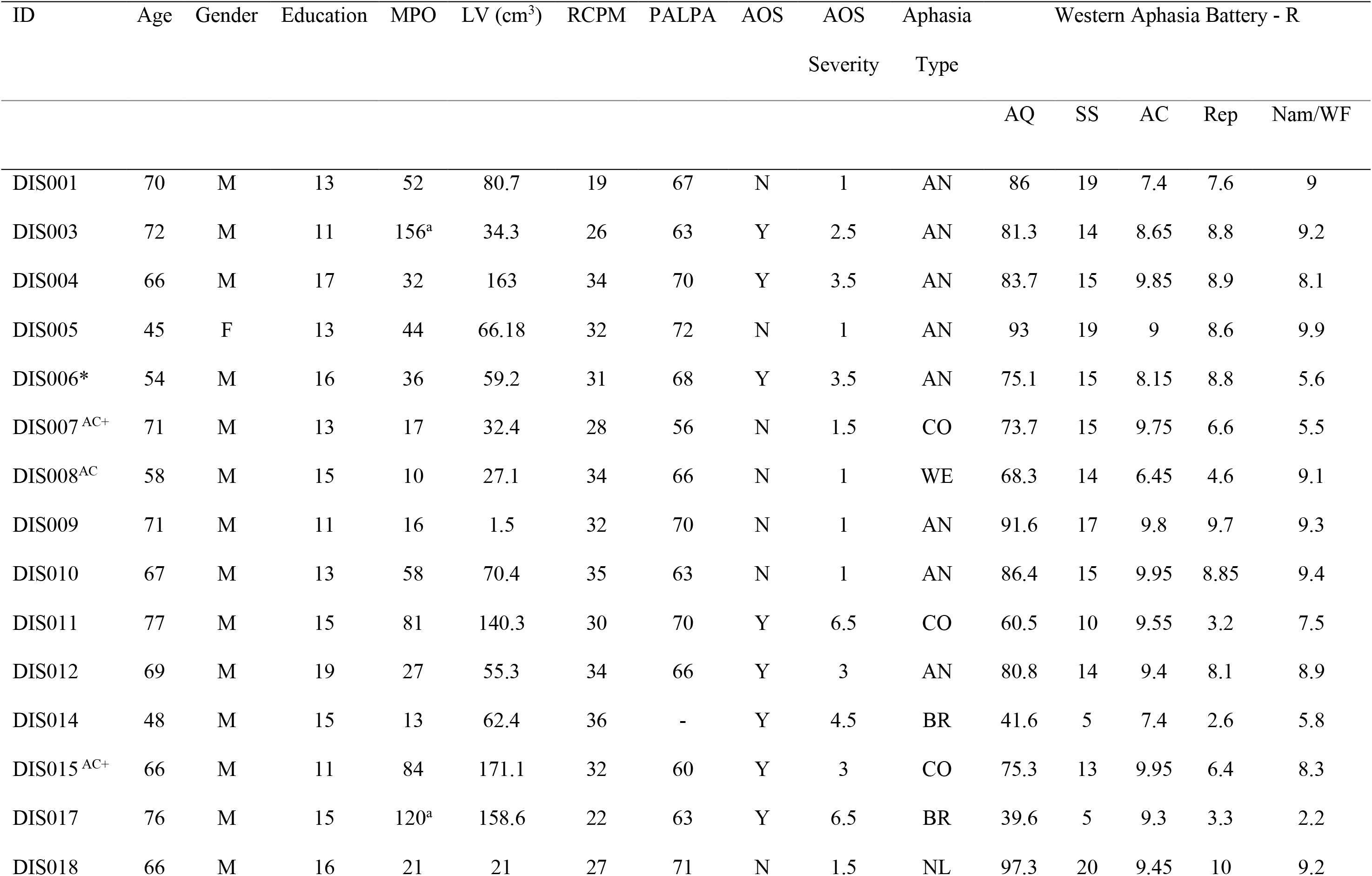
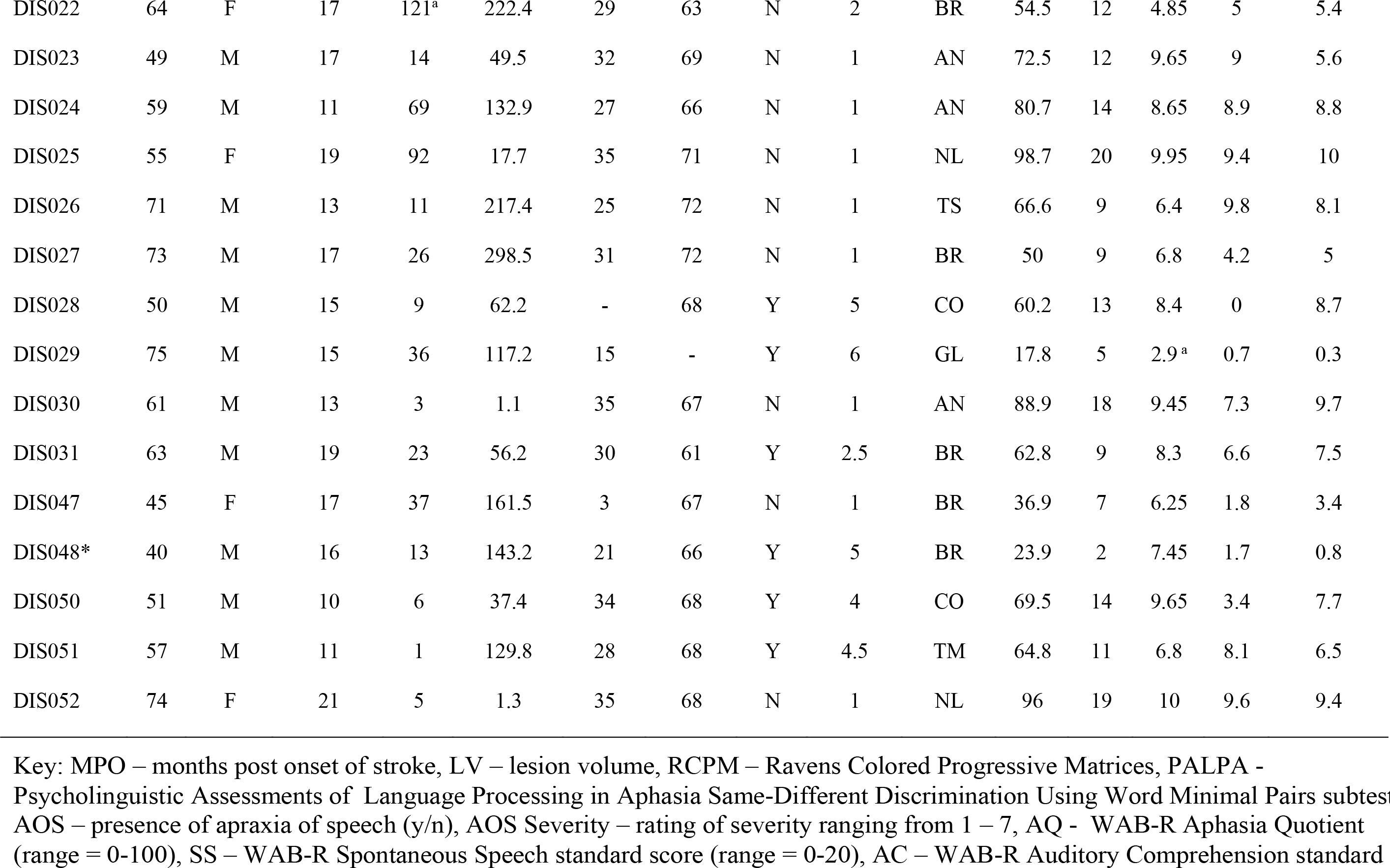
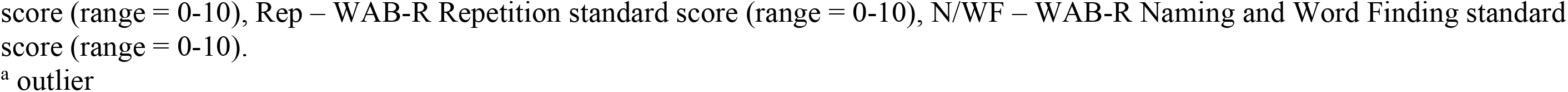
Demographic data and behavioral test scores for each participant with stroke. * participant’s primary language was not English. – indicates missing data.

**Table 2.**
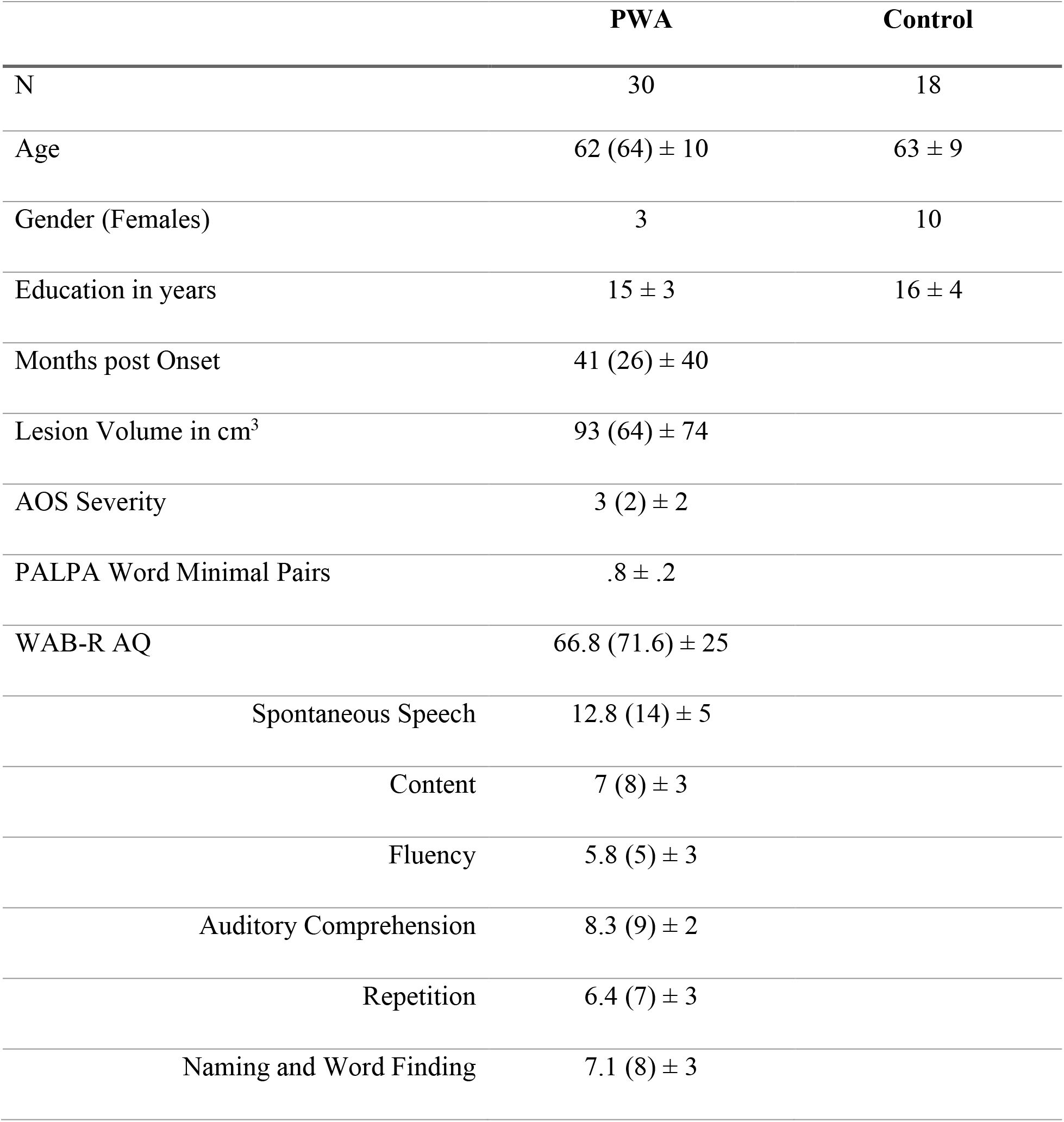
Means (medians) ± standard deviations for the group demographics for participants with aphasia and Control participants and the WAB-R Aphasia Quotient and subtest scaled scores. The groups did not differ significantly for age, *U* = 225.5, *p* = 0.84, or education, *U* = 285.5, *p* = 0.22. There were more females in the Control group than the PWA group, *X*^2^(1, *N* = 48) = 10, *p* = .002.

**Figure 1.**
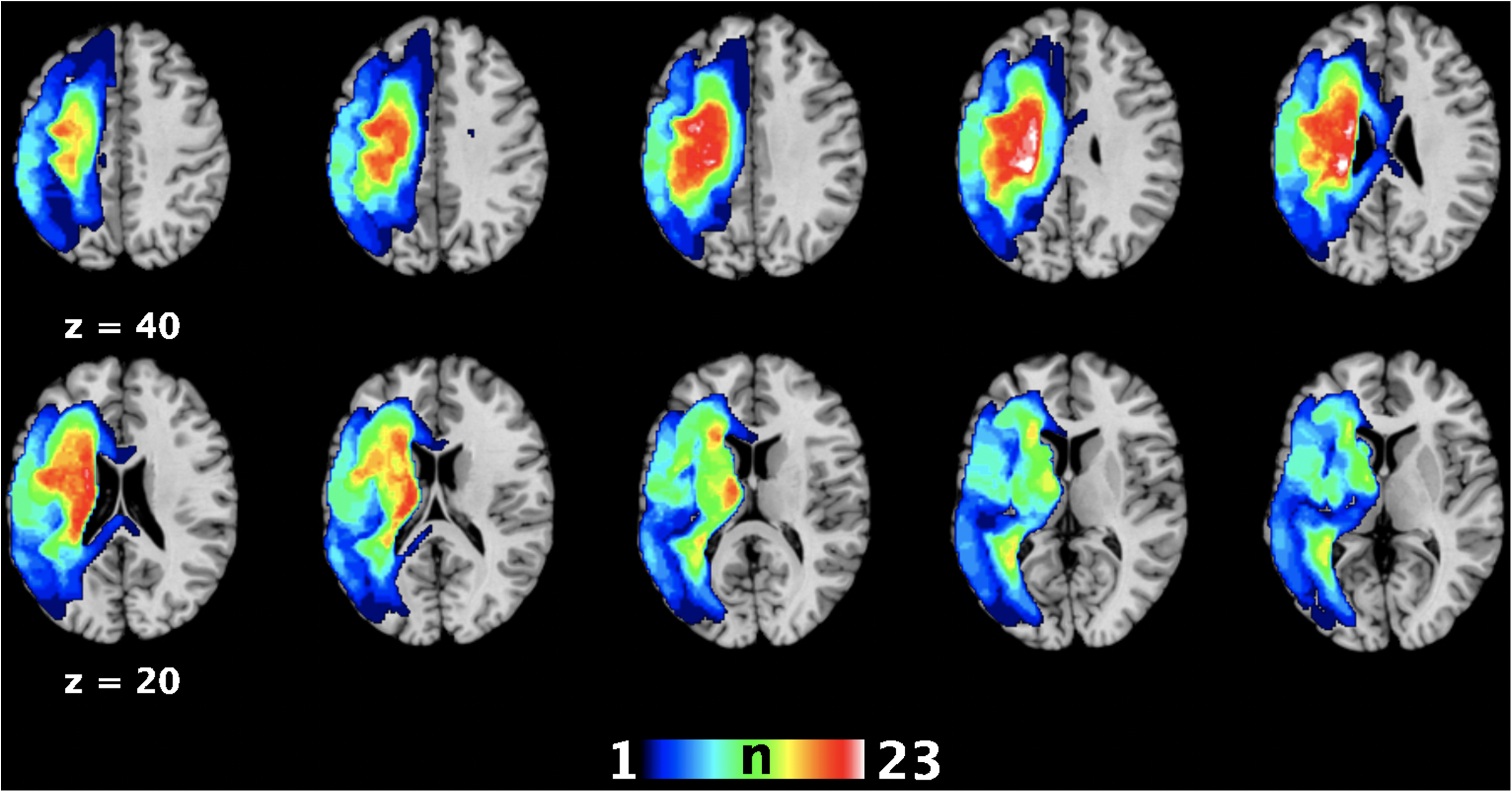
Lesion overlap map for all stroke participants demonstrating the extent of the lesion as well as the peak point of overlap amongst patients (in white). The peak overlap (23-24 of the PWAs had lesioned tissue) was in the left posterior insula (*x* = -24, *y* = -30, *z* = 27) and the body of the caudate (*x* = -21, *y* = -8, *z* = 26). Coordinates are in MNI space. Images generated in Mango (http://ric.uthscsa.edu/mango/mango.html, Lancaster et al., 2010).

PWAs with larger lesion volumes performed more poorly on WAB-R Spontaneous Speech, *r*(30) = -.67, *p* < .0001, Fluency, *r*(30) = -.67, *p* < .0001, Content, *r*(30) = -.66, *p* < .0001, and Naming and Word Finding, *r*(30) = -.58, *p* = .002, subtests. More severe AOS was associated with poorer performance on all WAB-R sub-tests (Spontaneous Speech, *r*(30) = -.55, *p* = .001; Fluency, *r*(30) = -.67, *p* < .0001; Content, *r*(30) = -.46, *p* = .018; Repetition, *r*(30) = - .65, *p* < .0001; and Naming and Word Finding, *r*(30) = -.60, *p* = .001), except AQ, *r*(30) = -.14, *p* = 0.49, and Auditory Comprehension, *r*(30) = -.20, *p* = .33. No such correlations existed between WAB-R scores and age, sex or months post onset of stroke (see Supplemental Table 1 for all correlations). Henceforth, connectivity amongst the regions will be evaluated in the stroke patients only and controlled for lesion volume and AOS severity.

### Image Acquisition

T1-weighted structural and resting state echo planar (EPI) functional MRI data were acquired on a Philips 3T TX MRI scanner. Blood-oxygen-level-dependent (BOLD) contrast [gradient-echo EPI pulse sequence, TR = 2.2 s, TE= 30 ms, flip angle = 90°, in-plane resolution=3.1 × 3.1 mm2, 36 axial slices (3.1mm thickness) covering the entire brain] was utilized to acquire 216 resting state echo planar images.

#### Image pre-processing

Structural scans were normalized to standard MNI space in SPM8 (http://www.fil.ion.ucl.ac.uk/spm) using the “unified segmentation” algorithm (Ashburner & Friston, 2005). For the stroke participants, an extra empirically derived tissue class (“lesion”) was added to the segmentation priors to be represented in a tissue class separate from gray/white/cerebrospinal fluid (Seghier, Ramlackhansingh, Crinion, Leff, & Price, 2008). All segmentation output images were smoothed with an isotropic kernel of 8 mm at full width at half maximum (FWHM). After smoothing, the value of each voxel in the image presented the probability that the tissue belongs to a single class and not to one of the others. The lesion tissue class image for each subject was also used to determine lesion volumes using the automated lesion identification algorithm (ALI toolbox) implemented in SPM8 (Seghier et al., 2008). This method is used in many analyses of stroke data, with established accuracy (Crinion et al., 2007). Though error in delineating the lesion is minimized with this and similar procedures (Pustina et al., 2016), there is potential for mischaracterization in periventricular lesions or when atrophy is significant. Lesion volumes (cm^3^) were calculated for each participant and included in the analyses described below.

EPI images were corrected for head movement by applying affine registration using a two-pass procedure in SPM8. Mean EPI images for each subject were created and spatially normalized to the MNI template as above and smoothed using a 5-mm FWHM Gaussian kernel. Variance associated with physiological noise and motion was removed to reduce false correlations (Bandettini & Bullmore, 2008; Jakobs et al., 2012). Data were then band-pass filtered, preserving frequencies between 0.01 and 0.08 Hz (Fox & Raichle, 2007; zu Eulenburg et al., 2012).

#### Region of interest selection

The brain regions used as regions of interest (ROIs) in our analysis are those proposed in the literature to be necessary for semantic or phonologic processing and for motor programming. Please see Table 3 for a list of ROIs and their coordinates, reasons for their selection as ROIs, and hypotheses about the WAB-R scores each may predict. Please also see Supplemental Figure 1 for a map of ROI locations. The bilateral homologs of all of the ROIs was also included in the study, given the known changes in laterality of language following stroke (Saur et al., 2006) and language-relevant function in the right hemisphere (Chai et al., 2016). Semantic ROIs were regions in which the structure (gray matter volume or density) was found previously to associate with tasks that isolated specific processes associated with access to word meaning. Several of the ROIs were borrowed from the work of Halai and colleagues (Halai et al., 2017). These investigators administered a large battery of language measures to a cohort of PWAs (*n* = 31) and factor analyzed the data, deriving factors identified as semantic, speech fluency, or phonology. Tasks loading on the semantic factor included confrontation naming, synonym judgment, spoken and written word-to-picture matching, and type-token ratio (Halai et al., 2017). Another study that contributed semantic ROIs investigated the correlations between gray matter density and change in word retrieval performance of PWAs on object or action picture naming tasks over time (recovery) (Hope et al., 2017). Finally, Seghier and colleagues isolated activation of the left ventral angular gyrus (AG) in processing of semantic matching, relative to perceptual matching, in healthy participants (Seghier et al., 2010).

**Table 3.**
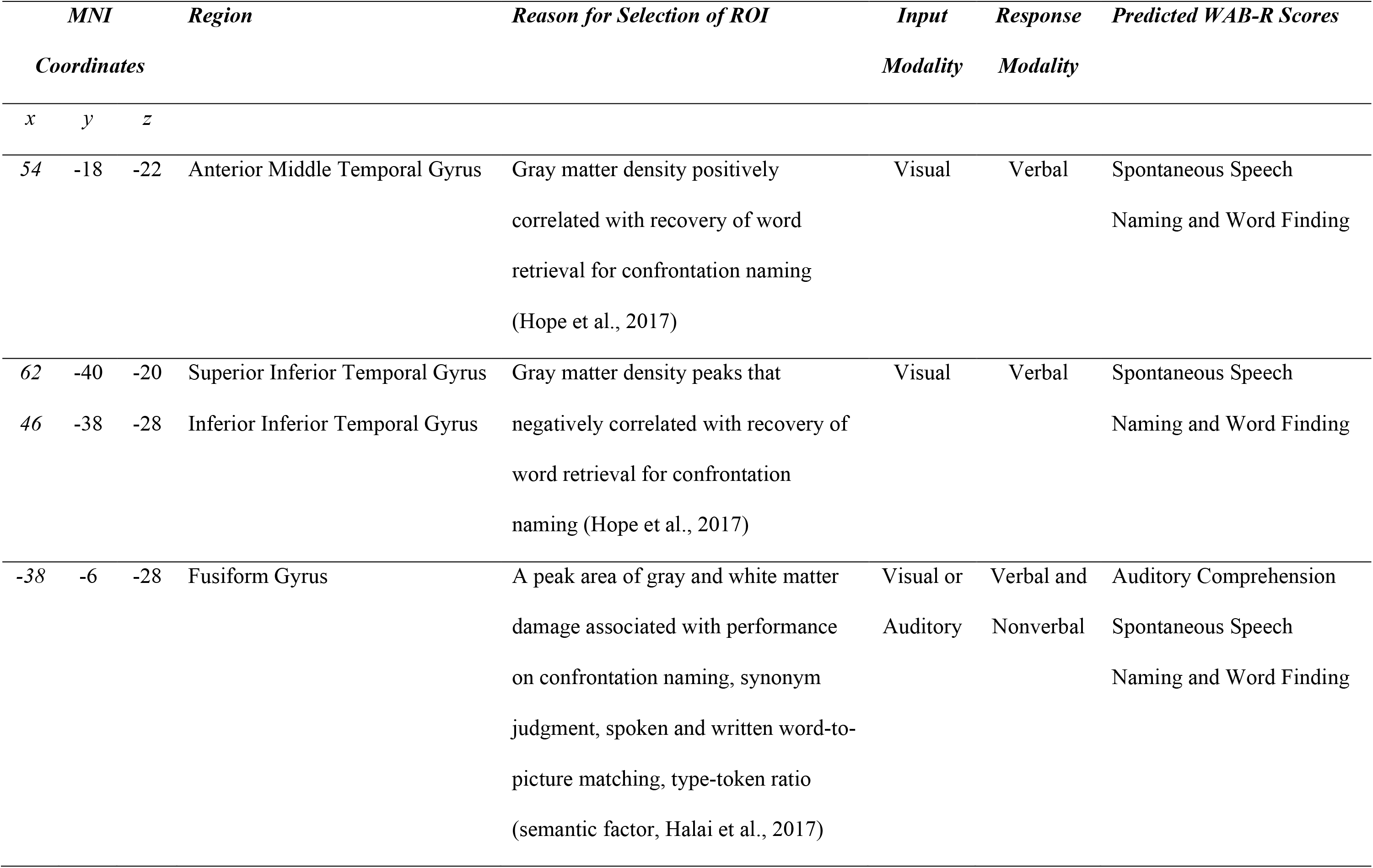
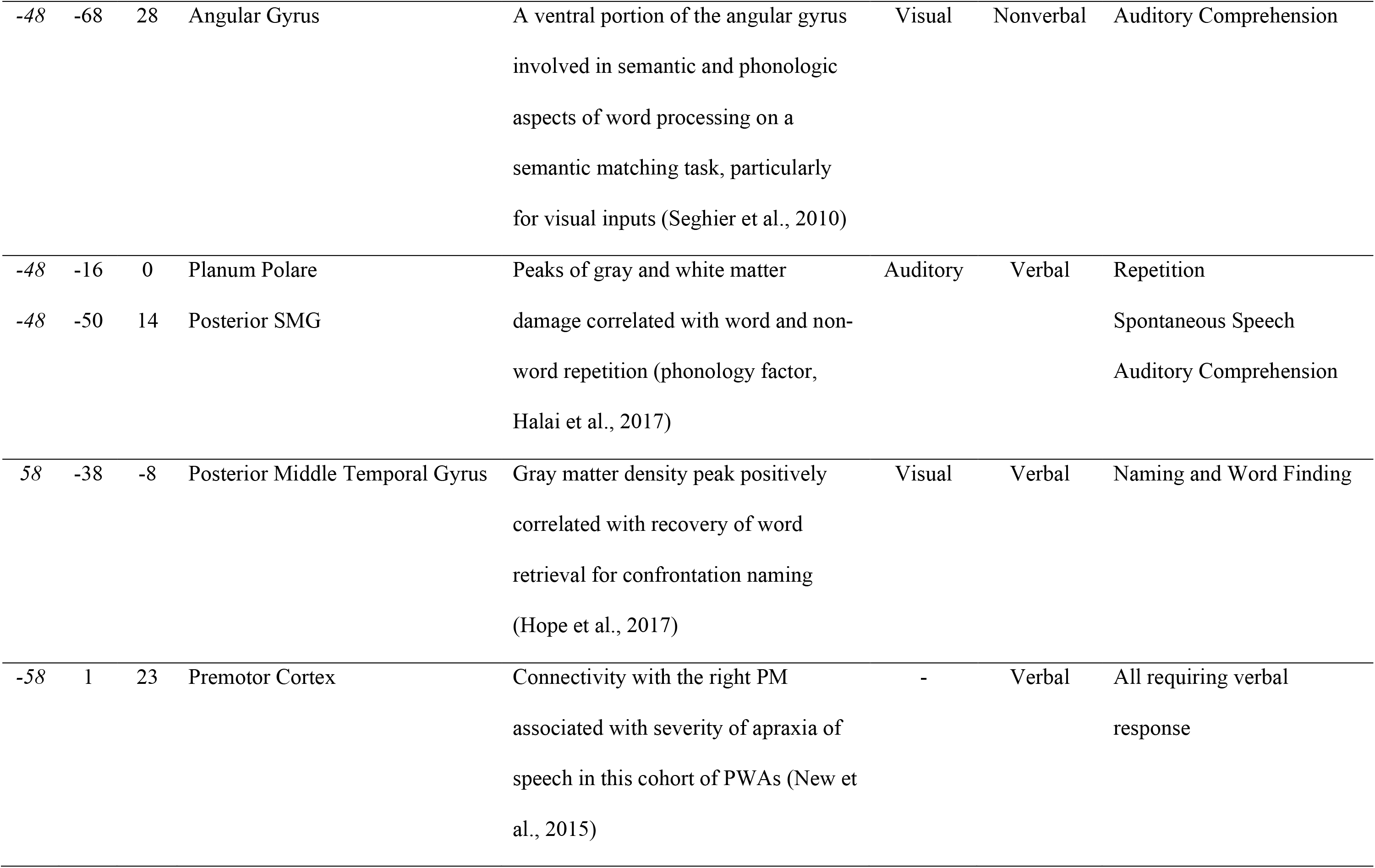
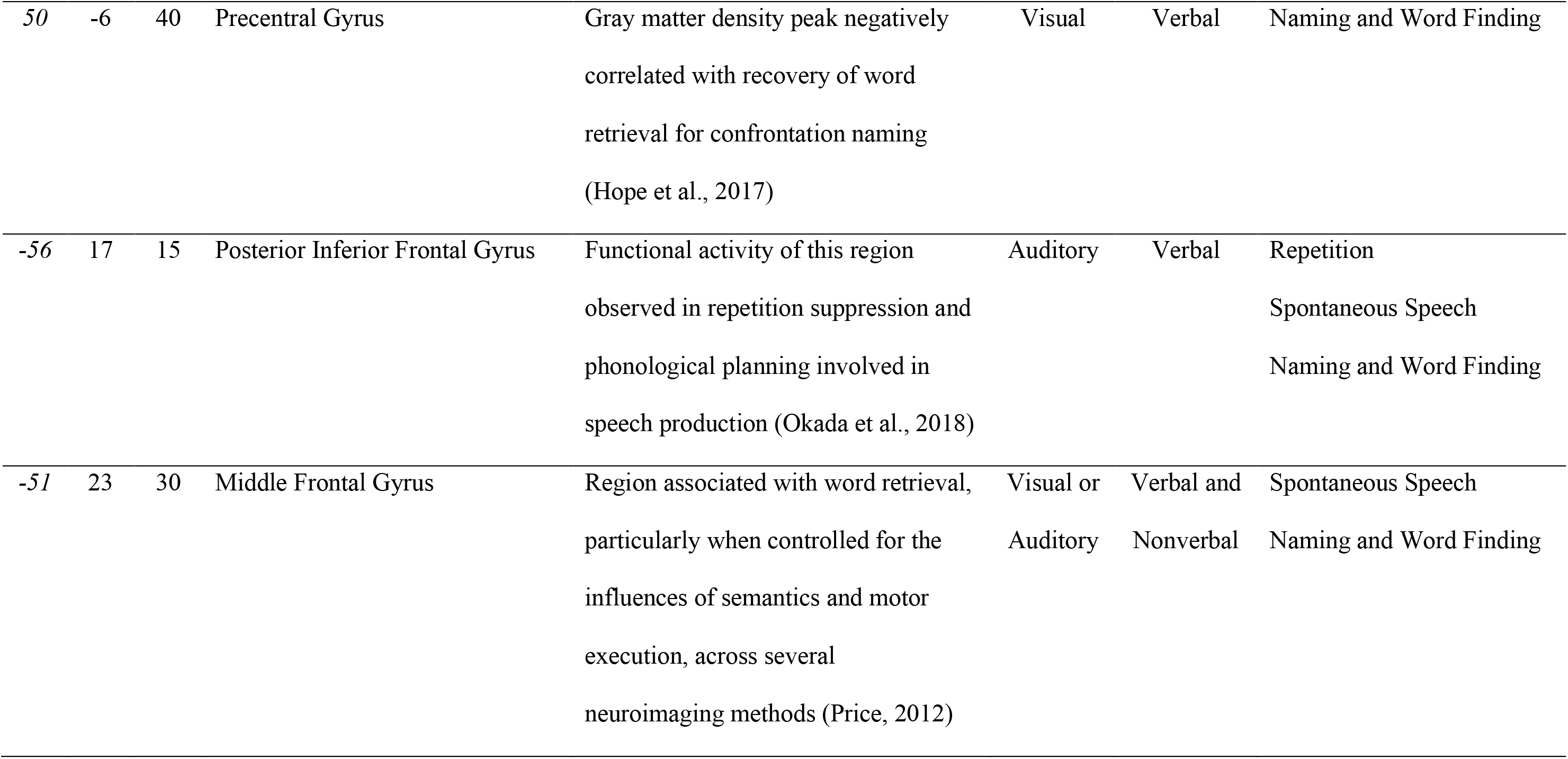
Coordinate locations for the regions of interest based on findings of either semantic, phonologic, or motor aspects of word retrieval or production, with input and response modality noted (input: A = auditory, V = visual; response: V = verbal, N = nonverbal). Regions labeled by coordinates provided by the authors and using the Montreal Neurological Institute’s (MNI) coordinate system in the Talairach Daemon (Lancaster et al., 2000).

ROIs attributed to phonologic aspects of word retrieval or production included: the left planum polare (PP) and left supramarginal gyrus (SMG) (phonologic factor from Halai et al., 2017), and left posterior inferior frontal gyrus (IFG) (Halai et al., 2017; Okada et al., 2018). The latter region was found to activate in an fMRI study of repetition suppression and the phonological planning involved in speech production (Okada et al., 2018). ROIs associated with speech-motor programming included the left premotor cortex (PM; New et al., 2015), found previously in this data to relate to AOS severity in this sample, and the right precentral gyrus (PcG)(Hope et al., 2017), noted by Hope and colleagues as the premotor cortex and found to negatively correlate with recovery of picture naming.

Having chosen these ROIs based on findings about their associations with fairly specific language task performance allowed for *a priori* hypotheses regarding relationships between functional connectivity amongst these regions and WAB-R performance (Table 3). For example, damage to the left fusiform gyrus was associated with poorer performance on tasks requiring recognition of speech and access to semantic representations (spoken word-to-picture matching; auditory input and nonverbal response), word retrieval for single-word production (confrontation naming) and fluent expression (type-token ratio) (Halai et al., 2017). As such, the left Fusiform is hypothesized to be connected with other ROIs involved in semantic processing for tasks with similar semantic access demands – i.e., Auditory Comprehension, Spontaneous Speech, and Naming and Word Finding.

#### Data analysis

The G*Power 3.1.9.4 software package was used for power calculations (Faul et al., 2007, 2009). The sample size of n = 48 provided 77% power to detect a large effect size, and 49% power to detect a medium effect size (Faul et al., 2009). Group differences for demographic variables and WAB-R scores were calculated in SPSS 24 (IBM Corporation, 2015). The distribution for each of the language variables was visually inspected on Q-Q plots and assessed using one-sample Kolmogorov-Smirnov tests, assessing the null hypothesis that the WAB-R variables are normally distributed. The null hypothesis was not rejected for the WAB-R AQ, SS, and Fluency, but rejected for the Naming and Word Finding (skewness = -1.13, kurtosis = 0.31, *D*(31) = .19, *p* = .005), Auditory Comprehension (skewness = -1.56, kurtosis = 2.39, *D*(31) = .18, *p* = .01) and Repetition (skewness = -.58, kurtosis = -1.10, *D*(31) = .18, *p* = .016) subtests, indicating that the latter were not normally distributed. To identify the influences of lesion volume or AOS severity on WAB-R scaled scores, Spearman correlations were computed.

#### Functional Connectivity

Each ROI’s time course was extracted within 5 mm of the respective peak coordinate (Table 1). Inter-ROI connectivity was assessed with linear, Pearson correlations (transformed into Fisher’s Z values) and independent samples t-tests determined group differences (*p* < .05 FDR corrected for multiple comparisons).

#### Generalized linear models

To identify potential predictors of WAB-R performance, the associations between FC and WAB-R scores were assessed with multivariate general linear models, with potential predictors being variables (including sex, lesion volume, months post onset and AOS severity) associated with each score at *p* < .05 with Type I or Type III sums of squares. The identified predictor variables for each WAB-R standard score were input to a generalized linear model. Generalized linear models (GLZs) provide a flexible extension of ordinary least squares regression and can accommodate dependent variables with either gaussian or non-gaussian error distributions (McCullagh and Nelder,1989). GLZs enable the analyst to specify a link function that relates the expected value of the response variable to the linear predictors in the model.

Visual inspection of the variable distributions and fit statistics indicated which model best fit the data (e.g., normal, Tweedie, Inverse Gaussian), and which predictors were significant (*p* < .05, corrected for multiple comparisons with the false discovery rate). Given the relatively small sample size, the robust estimator was used.

## Results

### Functional Connectivity

Figure 3A presents the 57 ROI-ROI resting-state connections that were coherent and significant in both the control and stroke participants. Of the 57, 36 were present in both groups, 16 only in the Controls, and 5 only in the PWAs (Figure 2, averages and standard deviations for each are presented in Supplemental Table 2). The presence of statistically significant connections in the PWA and Control groups suggests that these ROIs represent an intrinsically coherent network that is present at rest in healthy and lesioned brains. Controls had stronger FC than PWAs for the left and right PM connection, *t*(46) = 3.5, *p* < .05, FDR corrected.

**Figure 2.**
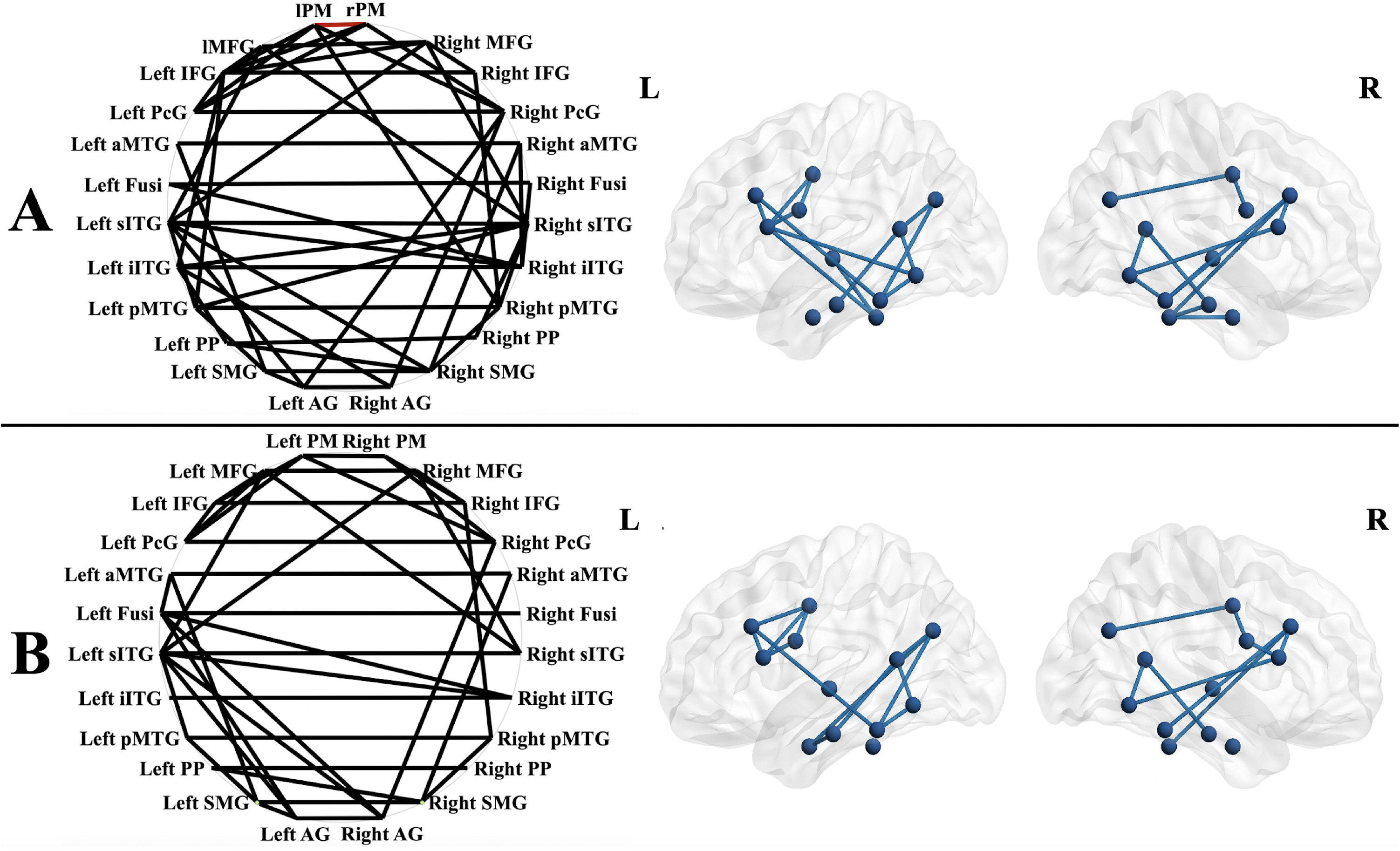
The HC group (A) had a greater number of resting state connections (indicated by black lines connecting regions of interest labeled) than the PWA group (B), but only differed significantly for strength of the left PM-right PM connection (HC > PWA). The circles (left) allow for visualization of all connections (between and within-hemisphere) in two dimensions, while the left and right lateral views (right) allow for visualization of within hemisphere connections and their anatomic locations. Figures generated in BrainNet Viewer (http://www.nitrc.org/projects/bnv/, Xia et al., 2013). Key: middle frontal gyrus (MFG), inferior frontal gyrus (IFG), precentral gyrus (PcG), planum polare (PP), fusiform gyrus (Fusi), anterior middle temporal gyrus (aMTG), superior aspect of the inferior temporal gyrus (sITG), inferior aspect of the inferior temporal gyrus (iITG), posterior middle temporal gyrus (pMTG), angular gyrus (AG), supramarginal gyrus (SMG).

**Figure 3.**
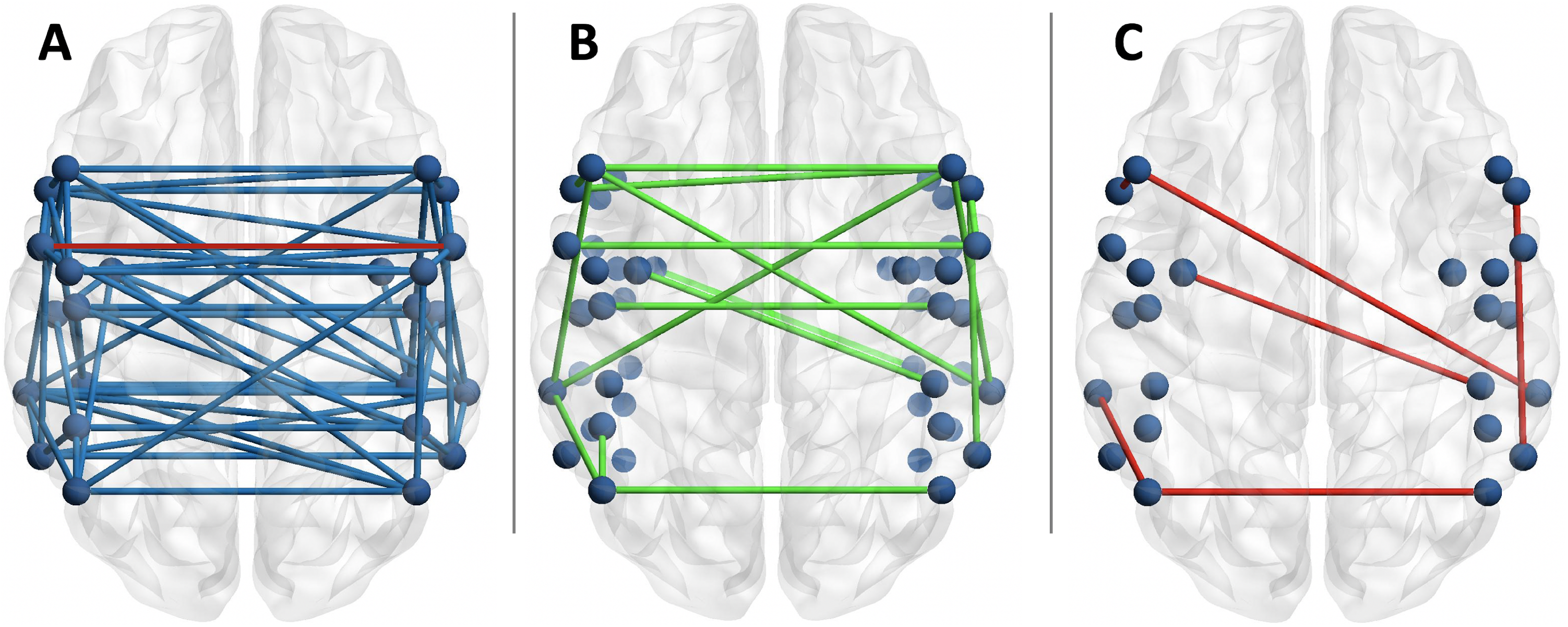
Of the connections present at rest in the study participants (A, red line indicates left PM-right PM group difference, *t*(46) = 3.5, *p* < .05 FDR corrected), 15 were potential predictors (green) of language performance on the WAB-R (B). Only 6 were significant predictors (red) of language performance in the generalized linear models (C).

Figure 3B presents the connections that were identified as potential predictors of one or more of the WAB-R scores, while Figure 3C presents those connections found to be significant predictors. For all of the generalized linear models, the β and Wald confidence intervals indicated that participant characteristics like age, lesion volume or AOS severity were more precise predictors of language scores with small differences in the scaled score (< 1 point), while significant FC predictors were less precise, with larger confidence intervals for scaled scores. Thus, for each WAB-R subtest, comparison of fit statistics (Akaike’s Information Criterion [AIC] and Bayesian Information Criterion [BIC], Müller et al., 2013) indicated whether the inclusion of the FC measures improved model fit. The results reported below represent the best fitting models for each score. Omnibus test statistics and Wald χ^2^ values and significance for all predictors by each WAB-R standard score are reported in Supplemental Table 3, best fitting model in Supplemental Table 4). Significant findings are noted in Table 4 and presented in Figure 4.

**Table 4.**
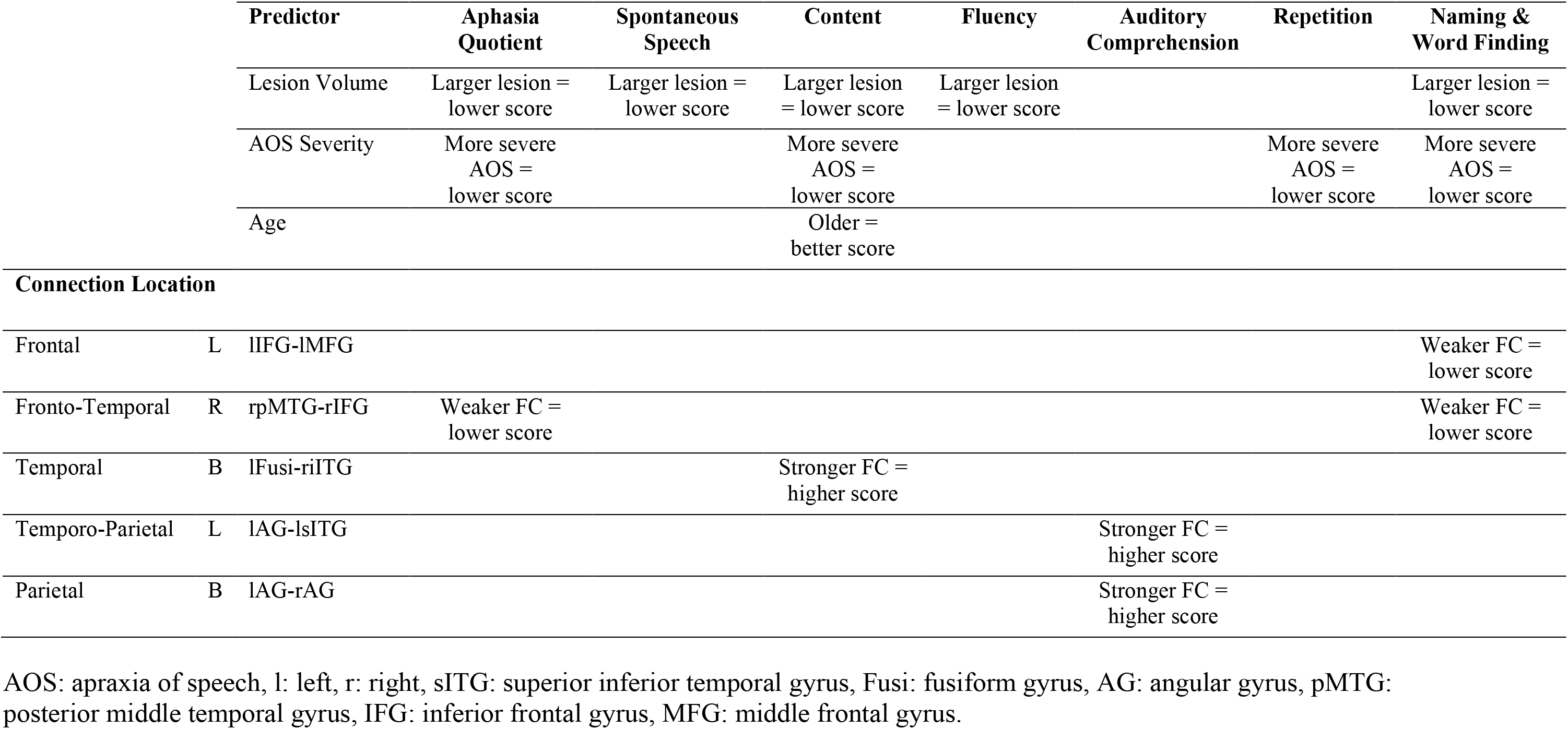
Significant predictors, all *X* ^2^ (df ranged from 2-4, *N* = 48) > 16, all *p*s < .05, FDR corrected for multiple comparisons, for each of the WAB-R standard scores included PWA characteristics including lesion volume, AOS severity, and age, as well as functional connectivity amongst ROIs in the frontal, temporal, and parietal lobes and involving unilateral (R – right, L – left) and bilateral connections (B).

**Table 5.** Summary of the predictors for each of the WAB scores. Based on the predictions of region-behavior associations predicted in Price (2012), and the findings in the present study, hypothesized functions for the connections are proposed. L – left hemisphere; R – right hemisphere; B – bilateral; + Positive predictor; - Negative predictor

|  |  |  |  | Western Aphasia Battery Scores |  |  |  |  |
| --- | --- | --- | --- | --- | --- | --- | --- | --- |
| Predictors |  |  |  | AQ | SS | AC | Rep | Naming |
| Lesion Volume |  |  |  | - | - |  |  | - |
| Apraxia of Speech Severity |  |  |  | - |  |  | - | - |
| Connection Location |  | Connection | Prediction about the Function of the Connection |  |  |  |  |  |
| Frontal | L | lIFG-lMFG | Phonological planning + word retrieval |  |  |  |  | - |
| Frontal-Temporal | R | rpMTG-rIFG | Naming recovery | - |  |  |  | - |
| Temporal | B | lAG-rAG | Semantic and phonological aspects of processing visual inputs |  |  | + |  |  |
| Temporal | L | lAG-lsITG | Naming recovery |  |  | + |  |  |

**Figure 4.**
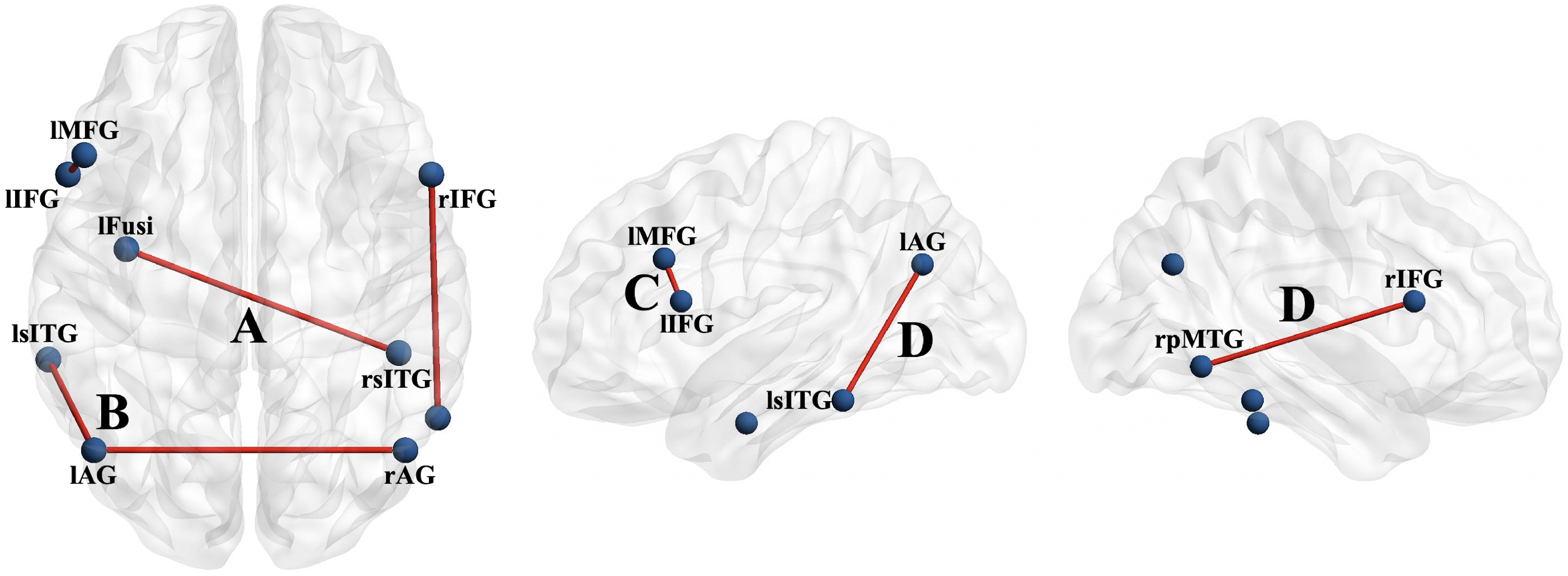
Axial (left), left hemisphere (middle), and right hemisphere (right) depictions of functional connectivity that is predictive of language performance on the WAB-R in the stroke participants. Table 4 provides the generalized linear models and the combinations of variables predicting performance, including each of the connections depicted here, all *X*^2^(df ranged from 2-4, *N* = 48) > 16, *p*s < .05 FDR corrected). Stronger connectivity strength between the left Fusiform and right inferior ITG (A) predicts better Content score. Stronger connectivity strength between the bilateral Angular Gyrus and between the left Angular Gyrus and left superior ITG (B) predicts better Auditory Comprehension score. Weaker connectivity strength between the left IFG and the left MFG (C) predicts better Naming and Word Findings score. Weaker connectivity strength between the right posterior MTG and right IFG (D) predicts better Aphasia Quotient and Naming and Word Finding scores. L – left, R – right. Figures generated in BrainNet Viewer (http://www.nitrc.org/projects/bnv/, Xia et al., 2013). Key: middle frontal gyrus (MFG), inferior frontal gyrus (IFG), fusiform gyrus (Fusi), superior aspect of the inferior temporal gyrus (sITG), inferior aspect of the inferior temporal gyrus (iITG), posterior middle temporal gyrus (pMTG), angular gyrus (AG).

### Prediction of WAB-R Aphasia Quotient

PWAs with smaller lesions, absence of or less severe AOS, and weaker functional connectivity between the right posterior middle temporal gyrus (pMTG) and right IFG had higher AQ scores.

#### Spontaneous Speech Standard Score

Lesion volume was the only significant predictor for WAB-R Spontaneous Speech (and Fluency) scores. However, model fit for the Content score, a component of the Spontaneous Speech standard score, indicated that older PWAs with smaller lesions, no or mild AOS, and stronger functional connectivity between left Fusiform and right inferior ITG had higher Content scores.

#### Auditory Comprehension Standard Score

Stronger FC between the left and right Angular Gyrus, and the left Angular Gyrus and left superior ITG predicted higher WAB-R Auditory Comprehension scores.

#### Repetition Standard Score

AOS severity was the only significant predictor of Repetition performance, with less severe AOS predicting higher Repetition scores.

#### Naming and Word Finding

Smaller lesion volumes, absent or minimal AOS, as well as stronger FC between the left IFG-left middle frontal gyrus (MFG) and the right pMTG-right IFG predicted higher WAB-R Naming and Word Finding scores.

## Discussion

The present study demonstrates that selecting semantic and phonologic regions of interest *a priori*, using existing neuroimaging findings from structural and functional studies of semantic and phonologic processing, elucidates a coherent resting-state network in healthy controls and individuals with aphasia secondary to left hemisphere stroke. This aligns with previous reports of coherent frequency oscillations amongst the regions of the language network at rest (e.g., Siegel et al., 2016). Controls had stronger connectivity than PWAs between the left and right premotor cortex, associated with the presence and severity of AOS (New et al., 2015), but there were no other significant group differences.

To validate this network of brain regions for its relevance to language, between-region functional connectivity along with participant characteristics known to be associated with aphasia severity (e.g., lesion volume, AOS severity), were tested as predictors of performance on a commonly used clinical assessment in aphasia, the WAB-R. Smaller lesion volume and presence of less severe AOS predicted better language performance on all the WAB-R subtests except Auditory Comprehension. In fact, Spontaneous Speech (particularly the Fluency rating) and Repetition were best predicted by these two variables – i.e., none of the FC variables improved model fit. However, the predictive models for the Aphasia Quotient, the Content component of Spontaneous Speech, and Naming and Word Finding scores fit best with the inclusion of FC variables between frontal, temporal and parietal regions in the left and right hemispheres that followed a pattern of stronger frontal and stronger temporo-parietal connectivity corresponding to better performance.

### Stronger connectivity involving frontal ROIs predicts better word retrieval

Stronger connectivity involving frontal lobe ROIs (left IFG-left MFG, right pMTG-right IFG), along with smaller lesion volumes and less severe AOS, predicted better Naming and Word Finding performance (Table 4). According to the studies from which these regions were borrowed, the left IFG (phonological planning for speech production, (Okada et al., 2018) and left MFG (word retrieval, Price, 2012) were predicted to be involved in concert for tasks requiring word retrieval and verbal expression – i.e., Spontaneous Speech (Content) and Naming and Word Finding. This prediction was correct for Naming and Word Finding, but not for Spontaneous Speech. However, similar connections involving the frontal lobes were *potential* but non-significant predictors for Spontaneous Speech (e.g., right pMTG-right IFG, see Supplemental Table 3) as well as its sub-components of Content (e.g., right superior ITG-right MFG) and Fluency (e.g., right superior ITG-left MFG). Nonetheless, this pattern of bilateral temporal-frontal connections highlights the importance of these relatively long-distance connections for verbal production of retrieved words in isolation (confrontation naming) and in connected language (picture description).

Weaker connectivity amongst frontal regions, particularly the left IFG-left MFG connection, may reflect an influence of the presence of AOS in 15 members of the PWA group. AOS severity has a negative impact on Naming and Word Finding performance (see Table 4), likely a result of the requirement of a verbal response. However, AOS severity was not correlated with left IFG-left MFG connectivity strength, *rho*(30) = .18, *p* = .34. These ROIs were hypothesized to be involved in phonological processing and word retrieval (see Table 3). Given the minimal effect of AOS on left IFG-left MFG connection strength, and the *a priori* predictions from the ROI selection (Table 3), the results suggest that this connection is likely involved in retrieving phonological representations of target words and translating phonological information for programming of motor execution.

The most robust finding in the study is the relationship between right pMTG-right IFG connectivity and Aphasia Quotient and Naming and Word Finding scores. Greater gray matter density of the right pMTG was found previously to be a positive predictor of naming recovery in individuals with aphasia – i.e., greater gray matter density of the right pMTG was associated with better naming and word finding performance (Hope et al., 2017). Hope and colleagues report that right hemisphere regions undergo systematic structural adaptations to support language functions, even in the chronic phase of recovery, and that the degree of these changes relates to word retrieval. The present study adds that weaker connectivity of the right pMTG-right IFG also associates with better AQ scores. It is outside the scope of the present study to identify whether or how right pMTG gray matter density may relate to right pMTG-right IFG connectivity, but we speculate that the structural adaptation of this structure may facilitate connections to other regions, and that FC of right pMTG-right IFG may be a maladaptive functional adaptation to aphasia. Longitudinal study of this connection would best address the validity and utility of this prediction. As well, having an understanding about whether and how the right pMTG-right IFG connection relates to language performance in healthy controls would aid in interpretation of this finding.

### Stronger connectivity amongst temporo-parietal regions predicts better access to semantics in visual or auditory modalities

The bilateral connection between the left Fusiform and right inferior ITG, two regions associated with access to semantic information for word retrieval, is necessary for imparting relevant Content in Spontaneous Speech. Both ROIs were predicted to be involved in semantic access for Spontaneous Speech and Naming and Word Finding, with the common factor being word retrieval. Both regions may be considered part of the “ventral stream”, or the “what” stream of Hickok and Poeppel (2007).

Additionally, stronger connectivity between the left Angular Gyrus-right Angular Gyrus and left Angular Gyrus-left superior ITG was the only significant predictor of Auditory Comprehension scores, i.e., no characteristics of aphasia or stroke predicted comprehension. These regions were found in previous studies to be involved in semantic association tasks (i.e., Angular Gyrus) and with recovery of word retrieval (i.e., superior ITG). The common denominator for these two tasks is access to word meaning, and thus we propose that these connections are necessary for access to semantics for understanding auditory input. These connections are also in agreement with the flow of language processing proposed in the Dual Stream Model (Hickok & Poeppel, 2007), with the Angular Gyrus aligning with the ‘conceptual network’ and in close proximity to the sensorimotor interface (sylvian-parieto-temporal area), and the superior ITG aligning with the lexical interface (linking phonological to semantic representations).

### Right hemisphere connectivity and language in aphasia

The right posterior MTG-right IFG connection would likely be structurally connected via a right hemisphere homolog of the ‘dorsal stream’ (Hickok & Poeppel, 2007). In the left hemisphere, it is conceivable that this connection could be strengthened (since weaker connectivity = more severe aphasia) with improvements in access to phonologic representations. This has been observed as improved naming and word finding performance following constraint-induced language therapy (CILT), via enhanced integrity of the left inferior longitudinal fasciculus (McKinnon et al., 2017). While McKinnon and colleagues found that group-based CILT improved naming performance and reduced semantic error production (likely ventral stream changes), others have reported improvement in other language measures (e.g., repetition in Bi et al., 2011, or WAB-R performance across subtests in Mozeiko et al., 2018). These data suggest that integrity and strength of frontal-temporal connections is predictive of language performance and *necessary* for language in aphasia. That is, input and output processing are integrated through this pathway and therefore treatments should affect change in it. However, the right hemisphere homolog of the dorsal stream has not yet been investigated as a target for change secondary to language interventions. Given the findings here, and those of Hope and colleagues (Hope et al., 2017), it is conceivable that structural and functional integrity of right hemisphere fronto-temporal connectivity may be a marker of outcomes in intervention.

The majority of studies in aphasia on left-right brain structure and function has operated under the theory that right hemisphere involvement in language processing is compensatory or maladaptive (Saur et al., 2006). Laterality of language function is known to shift to the right in the acute phase and re-shift to the left hemisphere with recovery (c.f., Hartwigsen & Saur, 2017), or to shift to the right with certain types of treatments (e.g., Melodic Intonation Therapy in Wan et al., 2014). The present study is merely a snapshot of resting-state functional connectivity in the chronic phase, and cannot address compensation, but the patterns of positive and negative predictive value for the bilateral connections suggest that right hemispheric connectivity is not simply compensatory or maladaptive. As mentioned above, aphasia severity is predicted, in part, by connectivity strength between the right pMTG-right IFG, and the significant bilateral predictors were positive – i.e., stronger connectivity = better performance. As such, strengthening these connections may be helpful in (1) improving semantic access for processing information regardless of the need for output (left Angular Gyrus-right Angular Gyrus predicting Auditory Comprehension) and (2) for formulating spontaneous speech (left Fusiform-right inferior ITG predicting Content in Spontaneous Speech).

### Importance of Bilateral Temporo-Parietal Connectivity

The bilateral connections that were predictive of language performance involved temporal or parietal regions (Figure 4). This finding is akin to those of Braun and colleagues (Braun et al., 2001) and Silbert and colleagues (Silbert et al., 2014) who report that earlier stages of lexical processing for comprehension and production occur in posterior regions bilaterally, while the later stages necessary for production are left-lateralized and more anterior. Similarly, a recent coordinate-based meta-analysis of sentence comprehension and production points to bilateral posterior temporal cortex (temporo-occipital cortex) in lexical-semantic processing (Walenski et al., 2019) for sentence production. All three of these studies were conducted in neurologically healthy, relatively young adults during sentence comprehension and/or production tasks but suggest roles of bilateral posterior temporal regions in lexical semantic processing for comprehension and production of complex language. Specific to resting-state functional connectivity, increased bilateral connectivity at rest is relevant to that observed during language processing (Tzourio-Mazoyer et al., 2015), and is predictive of better overall outcomes following stroke (Puig et al., 2018). We are not aware of other studies that have investigated *resting-state* connectivity in stroke and aphasia indicating anterior-posterior inter-hemispheric differences relative to language performance, though one other study has reported increased connectivity amongst posterior regions of the default mode network following a naming treatment (Marcotte et al., 2013). We therefore are cautious of over interpreting this finding, given that resting-state functional connectivity in stroke may be confounded by variation in hemodynamic lag (Siegel et al., 2017), and that the bulk of lesioned tissue in this cohort is in the left frontal cortex, thus replication in a larger study, potentially with hemodynamic lag as a variable, is warranted. Nonetheless, the data here suggest that inter-hemispheric connectivity involving the left Angular Gyrus and superior ITG is significant in predicting performance on language tasks requiring semantic and phonological access and maintenance of representations for comprehension and production, requiring synthesis of information processing across the language and other cognitive domains (Chai et al., 2016).

### A comment on compensatory connectivity

The clinical significance of the right pMTG-right IFG connection, potentially as a maladaptive response to stroke in the left hemisphere, is intriguing. To be clear, the data here suggest that the stronger the connectivity between these right hemisphere homologs of Broca’s and Wernicke’s areas, the poorer the language performance overall (stronger connectivity = more severe aphasia). It has been suggested that right hemisphere involvement in language processing in individuals with aphasia, as well as in healthy controls, may be indicative of “lexical learning” or resolution of lexical conflict in learning (Raboyeau et al., 2010). Though, in most cases, the right-sided activity during language performance, particularly in the right IFG, is sub-optimal (Rosen et al., 2000). Study of the whole brain at rest indicates that while the left hemisphere ROIs in this study may represent a *core* language network, right hemisphere homologs are considered to be *peripheral* with less stable (or more flexible) connectivity at rest, suggesting that it may have a less specialized functional role in language (Chai et al., 2016). For example, the right hemisphere auditory cortex (primarily the parieto-temporal junction) is involved in error detection and making feedback-feedforward predictions about verbal productions – i.e., when productions match the intended sounds (c.f., Houde et al., 2002; Kort et al., 2014). Findings of this effect have been observed in EEG or MEG specific to voicing (e.g., Parkinson et al., 2014) and vowel production (e.g., Niziolek et al., 2013) and at discrete time scales. However, it also has been documented as reduced right hemisphere MEG signal during whole-item versus fine-grained lexical decision analysis of phonological stimuli (pseudowords) – an effect opposite of that seen in the left hemisphere (Ylinen et al., 2015). Though these findings are not localized to the ROIs in this study per se, they document a specialized role of the right temporal cortex in monitoring verbal productions, which has also been noted in real-time monitoring of language production in individuals with aphasia (Sreedharan et al., 2019). Because this connection was a negative predictor of Naming and Word Finding here (weaker connection = better performance), which involves verbal production, we speculate that the right pMTG-right IFG connection may be involved in monitoring of verbal production in such a way that is maladaptive when uninhibited. However, the poor temporal resolution of fMRI relative to structural imaging or EEG/MEG data limits our ability to address this speculation in the present study.

### Limitations

Though the intent of using the WAB-R as the language measure allows for application to a well-known clinical measure for aphasia, it does not allow for specific definition of the language processes assessed in each subtest. The loosely defined language processes do not allow for more direct predictions about the many language processes involved in each subtest and potential disagreement about the hypotheses put forth in Table 4. Given that we had relatively low power to detect small-to-medium effect sizes, there may be a risk of Type II error. As such, replication of this study in a larger sample with more detailed characterization of the semantic-phonologic impairment, as well as extension to understanding whether and how rsfMRI relates to communication for life participation, is warranted. In addition, the intent and design of the study centered on a region of interest, seed-based analysis and did not consider the potential for use of differing time windows in the approach to account for variability in the hemodynamic response, or a lag of the response as a result of stroke. Future study may consider alternative approaches (e.g., as in (Chai et al., 2016)) that may not only control for hemodynamic lag, as well as consider the stability and functional specification of bilateral or right hemisphere connections. Finally, the cross-sectional design employed in this study precludes causal inferences, and a longitudinal study in future study would expand the utility of the predictions to recovery or treatment effects.

## Conclusions

The network of semantic and phonologic brain regions chosen as regions of interest for their roles in semantic and phonologic processing show patterns of functional connectivity that predict language performance in post-stroke aphasia. Connections between regions that relate to recovery of specific language functions – e.g., naming (Hope et al., 2017), were most predictive of language performance. For example, at least one of the ROIs drawn from the Hope study was present in 3 of the 5 connections found to predict language performance. Several of these connections involve the right hemisphere, and bilateral connections were limited to the temporal cortex, supporting the notion that semantic processing and lexical access involves a bilateral posterior subnetwork in individuals with aphasia. Other connections involved frontal regions, and were largely left lateralized, with the exception of the right pMTG-right IFG which was predictive of aphasia severity and word retrieval for verbal production.

The connections that were predictive of language performance in aphasia have potential as targets for the study of neuroplastic functional adaptations as evidence of treatment efficacy. For example, one might predict that efficacy of treatments targeting semantic access for word retrieval (e.g., Semantic Feature Analysis; Boyle & Coehlo, 1995) should increase functional connectivity between the left Angular Gyrus-left superior ITG and left Fusiform-right inferior ITG, and weaker connectivity between the right posterior MTG-right IFG. Or, one might predict that connectivity of the left IFG-left MFG should strengthen given a phonologically-based treatment (e.g., Phonological Components Analysis; Leonard, Rochon, & Laird, 2008). Further study is required to determine the utility of such predictions. Rather, this study demonstrates that differing types of information may be gained by careful consideration of the brain regions assessed and fine-tuning the analytic approach to identify specific language processes, even when using clinical assessment measures. These types of data may be invaluable in determining whether the changes that clinicians seek through intervention are (1) targeting the correct neural system to address a language process, and (2) guiding outcomes to address treatment effects. Future study is needed to determine whether the FC targets identified here can be useful in making these determinations.

## Data Availability

The data are not currently available, but may be requested from the first author.

## Acknowledgements

The authors acknowledge the contributions of members of the University of New Hampshire Cognition, Brain and Language Team (CoBALT) Hannah Franz, Gwyneth Horne, Mara Callahan, Jessica Lee and Elizabeth Kinney for their help in organizing and analyzing the data. They also thank Cathy Price and the Predicting Language Outcomes and Recovery after Stroke (PLORAS) laboratory for conducting the automated lesion identification (ALI) for this project. This work was supported by the National Health and Medical Research Council project grant 632763 and Australian Research Council Future Fellowship FT120100355 (Principal Investigator: Kirrie J. Ballard, Ph.D.).

## Notes

Funding: This work was supported by the National Health and Medical Research Council project grant 632763 and Australian Research Council Future Fellowship FT120100355 (Principal Investigator: Kirrie J. Ballard, Ph.D.).

### Competing Interest Statement

The authors have declared no competing interest.

### Author Declarations

All procedures conformed to the Declaration of Helsinki (BMJ 1991; 302: 1194) and were approved by the Sydney Local Health District Human Research Ethics Committee, Concord Repatriation General Hospital in Sydney and the University of Sydney, Australia.

